# A retrospective analysis of the diagnostic performance of an FDA approved software for the detection of intracranial hemorrhage

**DOI:** 10.1101/2023.11.02.23297974

**Authors:** Bianca Pourmussa, David Gorovoy

## Abstract

**Objective:** To determine the sensitivity, specificity, accuracy, positive predictive value (PPV), and negative predictive value (NPV) of Rapid ICH, a commercially available AI model, in detecting intracranial hemorrhage (ICH) on non-contrast computed tomography (NCCT) examinations of the head at a single regional medical center.

**Methods:** RapidAI’s Rapid ICH is incorporated into real time hospital workflow to assist radiologists in the identification of ICH on NCCT examinations of the head. 412 examinations from August 2022 to January 2023 were pulled for analysis. Scans in which it was unclear if ICH was present or not, as well as scans significantly affected by motion artifact were excluded from the study. The sensitivity, specificity, accuracy, PPV, and NPV of the software were then assessed retrospectively for the remaining 406 NCCT examinations using prior radiologist report as the ground-truth. A two tailed *z* test with α = 0.05 was preformed to determine if the sensitivity and specificity of the software in this study were significantly different from Rapid ICH’s reported sensitivity and specificity. Additionally, the software’s performance was analyzed separately for the male and female populations and a chi-square test of independence was used to determine if model correctness significantly depended on sex.

**Results:** Of the 406 scans assessed, Rapid ICH flagged 82 ICH positive cases and 324 ICH negative cases. There were 80 examinations (19.7%) truly positive for ICH and 326 examinations (80.3%) negative for ICH. This resulted in a sensitivity of 71.3%, 95% CI [61.3%-81.2%], a specificity of 92.3%, 95% CI [89.4%-95.2%], an accuracy of 88.2%, 95% CI [85.0%-91.3%], a PPV of 69.5%, 95% CI [59.5%-79.5%], and an NPV of 92.9%, 95% CI [90.1%-95.7%]. Two examinations were excluded due to no existing information on patient sex in the electronic medical record. The resulting sensitivity was significantly different from the sensitivity reported by Rapid ICH (95%), *z* = 2.60, *p* = .009 although the resulting specificity was not significantly different from the specificity reported by Rapid ICH (94%), *z* = 0.65, *p* = .517. The model performance did not depend on sex per the chi-square test of independence: *X*^*2*^ (1 degree of freedom, *N* = 404) = 1.95, *p* = .162 (*p* > 0.05).

**Conclusion:** Rapid ICH demonstrates exceptional capability in the identification of ICH, but its performance when used at this site differs from the values advertised by the company, and from assessments of the model’s performance by other research groups. Specifically, the sensitivity of the software at this site is significantly different from the sensitivity reported by the company. These results underscore the necessity for independent evaluation of the software at institutions where it is implemented.

## Introduction

Artificial intelligence (AI) is becoming increasingly popular for the analysis of diagnostically pertinent data in healthcare. Radiology has been particularly affected by this shift; algorithms are used for the automated detection of pathological findings on medical imaging. It is predicted that the medical imaging machine learning market will hit $2 billion by 2023 (Harris, 2019). This robust financial statistic reflects the demand for efficient, systematized, and accurate medical imaging machine learning algorithms.

This demand for medical imaging software emerges within the context of a chronic shortage of radiologists in the US where COVID-19 has instigated unprecedented resignation rates (Reeves, 2022). Radiology had been disproportionality affected by resource shortages, even prior to the COVID-19 pandemic; with the development of more complex imaging equipment and increasing reliance on scans for diagnosis, scan volume has increased. Additionally, the routine inclusion of double reading within hospital workflow potentiates this issue (Bruls & Kwee, 2020; Rao et al., 2021) and there is escalating concern about burnout in the field. The threat of burnout induced medical errors (Ruutiainen et al., 2013) within this resource strained environment accentuates the applicability of AI for medical imaging.

There exist several FDA approved AI models (Chen et al., 2021), some of which are already in use by healthcare centers. Some of these commercially available products are specifically used for the detection of intracranial hemorrhage (ICH) on non-contrast computed tomography (NCCT) examinations of the head. Early diagnosis of ICH is crucial for optimizing patient outcomes (Elliott & Smith, 2010) and this software can rapidly detect bleeds, creating a streamlined triage process that decreases the time between presentation and treatment while promising to reduce the resource strain on healthcare centers and physicians.

Although AI has the potential to optimize the reading of medical imaging, there is evidence to suggest that these models are not completely generalizable and do not identify ICH with the same proficiency across different sites. One group evaluated AIDoc, a commercially approved deep learning software, in the detection of ICH (Kau et al., 2022). They found an accuracy of 94%, a marked decrease from a different study reporting an accuracy of 98% (Ojeda et al., 2019). Kau and colleagues also noted that residents and radiologists under time pressure significantly outperformed the model.

These discordant results introduce the fundamental question of generalizability. It is particularly important to explore this question with rigor given that FDA approval of these algorithms only requires internal review, not peer review (Tariq et al., 2020), thus generalizability is not sufficiently accounted for in model validation. What is required is comprehensive evaluation of the performance of these models in various clinical settings so that their limits are entirely understood.

Another commercially available, FDA approved model distributed by RapidAI, Rapid ICH, advertises a sensitivity of 95.0% and specificity of 94.0%(RapidAI, 2020). We aim to assess the sensitivity, specificity, and accuracy of Rapid ICH, when utilized for the detection of ICH at a single regional hospital center site. In this retrospective analysis, we intend to better understand the generalizability of this model so that it may be appropriately applied within the clinical environment to systematically optimize hospital workflow and ultimately improve patient outcomes.

## Materials & Methods

This retrospective, single-center study was approved by Pearl IRB, an independent institutional review board. Permission to access patient data was granted by the institutional review board. The institutional review board waived the requirement for informed consent given the retrospective nature of this study.

### Patient Population

Inpatient and emergency room NCCT examinations of the head preformed between August 2022 and January 2023, on patients between the ages of 11 to >=89 (patient ages greater than 89 were masked per HIPAA regulations), were included in this study. Patients with an age >= 89 were treated as having an age of 89. Scans were obtained from a single regional medical center and represent patients with a variety of clinical presentations. Both repeat acquisitions and initial scans were included in the study, so multiple scans from different time points for a singular patient existed in this image cohort. Scans with significant motion artifact, even in the case that motion most likely had an impact on the AI software output, were included unless there existed a repeat acquisition with no motion impact. In this case, the repeated examination was used in place of the motion impacted examination. Ambiguous scans were reviewed by a board-certified radiologist. If the determination of positive or negative ICH remained unclear so that the ground truth could not be determined, the scan was excluded from the final analyses (Figure 1).

**Figure I.**
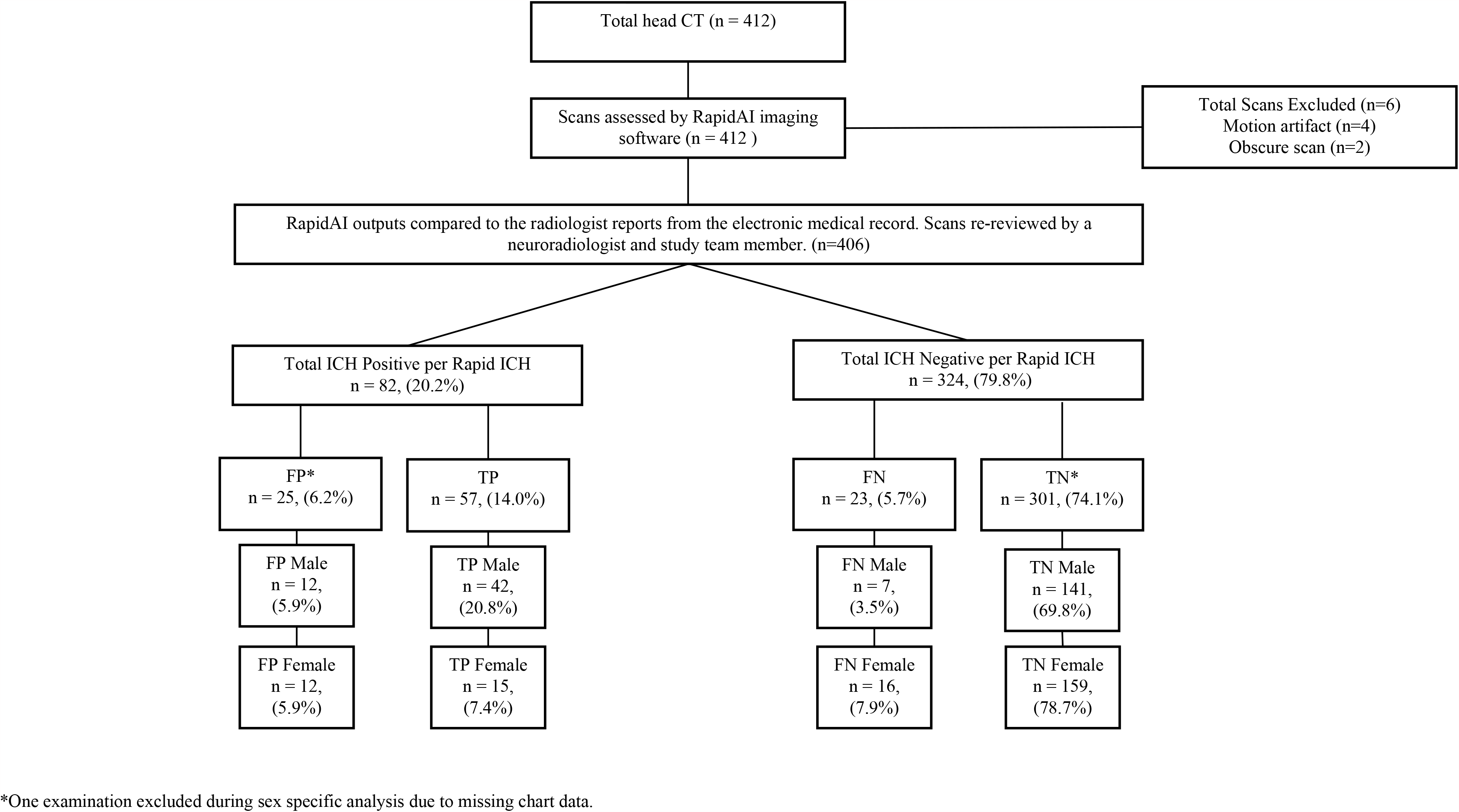
General study workflow and primary results of analysis of software output versus ground truth as defined by the radiologist *One examination excluded during sex specific analysis due to missing chart data.

### Image Acquisition

NCCT imaging was performed on GE Lightspeed volumetric computed tomography (VCT) 64 slice scanner with version vct_zeta.4 software. Scans were acquired under the following parameters: single collimation width, 0.625 mm; pixel dimension of image, 512x512; tube voltage, 120 kVP; adaptive tube current, 269 mA; pixel spacing, 0.488281/0.488281; windowing width/level, 80/40; convolution kernel, standard; rotation direction, counterclockwise; revolution time, 1 s; reconstruction diameter, 250 mm. Radiologists would have manually altered window settings during image review if necessary.

### AI Software: RapidAI

The scans were analyzed by the commercially available software, RapidAI, for the presence of ICH. RapidAI is an FDA approved software developed to automatically detect and flag ICH positive scans. According to the company’s literature, the model was trained on images representing a diverse array of pathologies from US and Australian patients, on multiple scanner types. The ground truth in the training data set was defined by a radiologist. Clinical validation was performed on a separate dataset from patients from US and Brazil. The company states that following adjudication by radiologists, the model had a sensitivity of 95% and specificity of 94% for the detection of ICH (RapidAI, 2020). Following their most recent FDA clearance in November 2022, RapidAI currently advertises a sensitivity of 98.1%, specificity of 99.7% for the detection of ICH, the highest sensitivity and specificity on the market for commercially available ICH detection products, with the ability to detect bleeds as small as 0.4 ml (*Rapid ICH Receives New FDA Clearance with Highest Sensitivity and Specificity on the Market*, 2022; RapidAI, 2023). In the current study, the older software version with a sensitivity and specificity of 95.0% and 94.0% was used since the site had not upgraded at the time of image analysis. The software is configured to detect ICH subtypes including intraparenchymal (IPH), intraventricular (IVH), subdural (SDH), and subarachnoid (SAH) but is not trained to detect hemorrhagic transformation.

412 NCCT examinations of the head with Rapid interpretations were retrospectively reviewed. Rapid outputs were contained in a separate series on the study. The outputs of the software were compared to the physician notes which were used to establish the ground-truth. A model error rate was then generated. The scans and ground truth labels were secondarily reviewed by the authors, including a board-certified radiologist. The software outputs were manually labelled as false positive (FP), true positive (TP), true negative (TN), and false negative (FN). Patient sex was obtained from the electronic medical record. Two patients in the sex specific analysis were excluded because of missing information in the electronic medical record.

### Statistical Analysis

Statistical analyses were run on version 16.69.1 of Microsoft Excel and version 6.5.4 of Jupyter Notebook, a computing platform used with version 3.11.5 of Python. The SciPy library, a free and open-source Python library, was used for computations. The sensitivity and specificity of the software in this study were compared to the sensitivity and specificity published by Rapid using a two-tailed Z test. A sampling distribution of sensitivity and specificity was generated using 100,000 random samples of 100 batches of outcomes (TP, FP, TN, and FN) from the dataset to generate a standard deviation for the population. The observed sensitivity and specificity and the standard deviation derived from random sampling were compared to the expected sensitivity of 95% and specificity of 94% published by Rapid to produce the *z* score and *p* value with α = 0.05. Sensitivity, specificity, accuracy, positive predictive value (PPV), and negative predictive value (NPV) were calculated for the entire set of cases and for females and males separately. 95% confidence intervals were then calculated for these statistics. Additionally, a chi-square test of independence was performed to determine if there is a statistically significant association between model performance and sex since a previous group (Voter et al., 2021) found that model performance was influenced by demographic factors.

## Results

A total of 412 NCCT examinations of the head were analyzed by the RapidAI software, and 6 scans were excluded prior to the final analyses due to motion artifact or unclear diagnosis (Figure I). 82 (20.2%) scans were flagged as positive for ICH and 324 (79.8%) scans were flagged as ICH negative by the software (Figure I). There were actually 80 examinations (19.7%) with true ICH and 326 examinations (80.3%) without ICH. Scans represented a patient population of 202 (50%) females and 202 (50%) males with a mean female age of 66.6 and mean male age of 60.3. ICH was truly present in 15.3% of females and 24.3% of males (Table II).

Of the software outputs, 25 (6.2%) were FP and 57 (14.0%) were TP (Figure I). The image features contributing to the FP results are described in Table III. FP outputs were a result of various features, but motion and residual contrast were the primary identifiable contributors. Additionally, 4 examinations triggered a positive output by the software in the absence of a bleed with no identifiable artifact noted on the scan.

Of the 324 cases flagged as ICH negative by the software, 23 (5.7%) were FN and 301 (74.1%) were TN (Figure I). FN cases were parsed by bleed type; 7 (30.4%) of FN outputs represented an intra-axial bleed (IPH), while 15 (65.2%) of these outputs represented an extra-axial bleed (SDH, SAH, or EH). One bleed type (4.3%) was unclear from the scan (Figure II).

**Figure 2.**
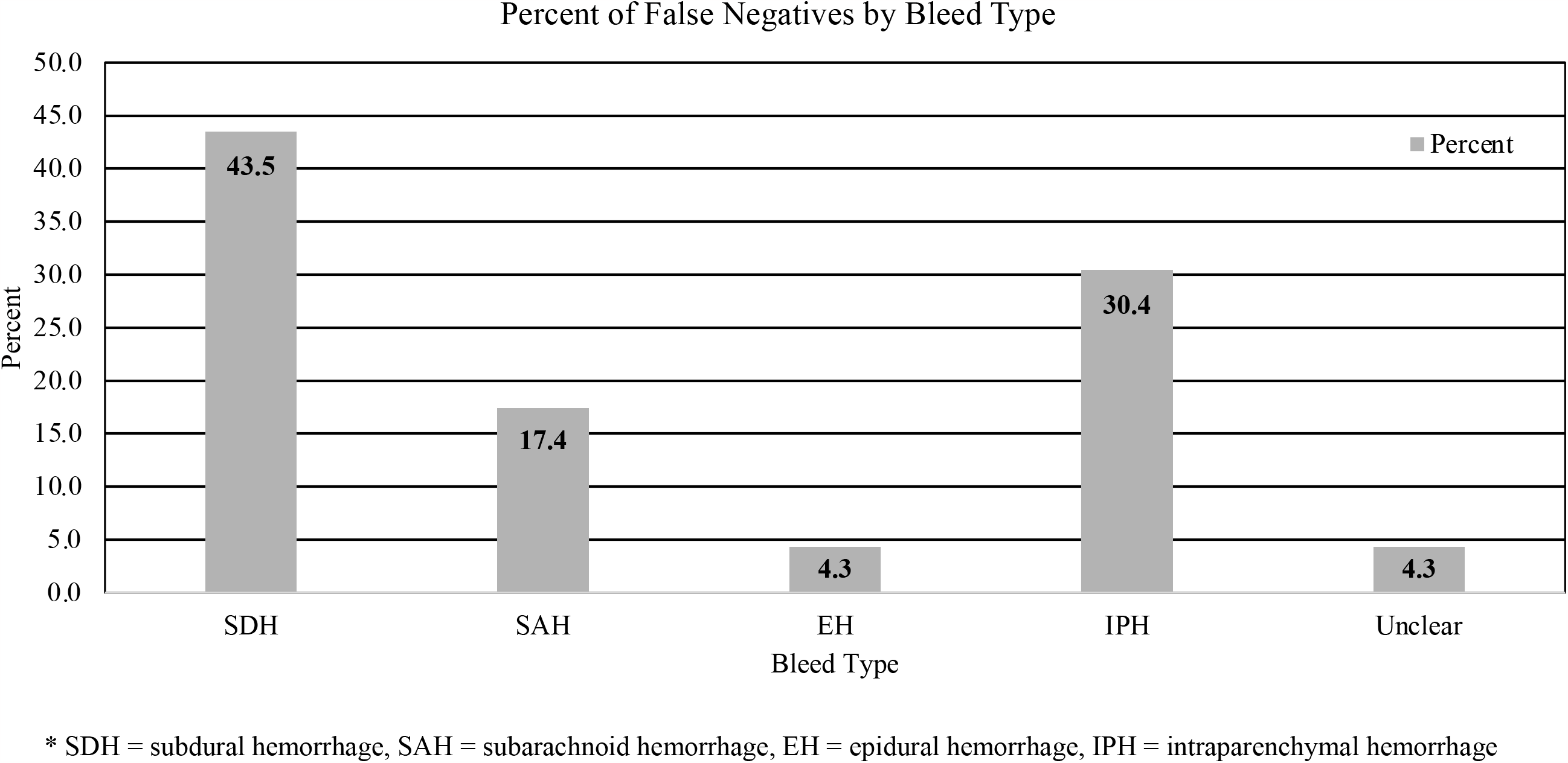
Frequency of false negative outputs per bleed type* * SDH = subdural hemorrhage, SAH = subarachnoid hemorrhage, EH = epidural hemorrhage, IPH = intraparenchymal hemorrhage

The sensitivity of the software was 71.3%, 95% CI [61.3%-81.2%], the specificity was 92.3%, 95% CI [89.4%-95.2%], and the accuracy was 88.2%, 95% CI [85.0%-91.3%]. Additionally, it determined that the PPV was 69.5%, 95% CI [59.5%-79.5%], and the NPV was 92.9%, 95% CI [90.1%-95.7%] (Table I). The resulting sensitivity was significantly different from the sensitivity reported by Rapid ICH (95%), *z* = -2.60, *p* = .009 but the observed specificity was not significantly different from the specificity reported by Rapid ICH (94%), *z* = 0.65, *p* = .517.

**Table I.** Performance parameters of Rapid ICH when applied to all examinations and for males and females separately.

| Statistic | Overall (% , [95% CI]) | Female (% , [95% CI]) | Male (% , [95% CI]) |
| --- | --- | --- | --- |
| Sensitivity | 71.3, [61.3-81.2] | 48.4, [30.8-66.0] | 85.7, [75.9-95.5] |
| Specificity | 92.3, [89.4-95.2] | 93.0, [89.2-96.8] | 92.2, [87.9-96.4] |
| Accuracy | 88.2, [85.0-91.3] | 86.1, [81.4-90.9] | 90.6, [86.6-94.6] |
| Positive Predictive Value | 69.5, [59.5-79.5] | 55.6, [36.8-74.3] | 77.8, [66.7-88.9] |
| Negative Predictive Value | 92.9, [90.1-95.7] | 90.9, [86.6-95.1] | 95.3, [91.9-98.7] |

**Table II.** Population Characteristics: Sex, true counts of bleeds, and age.

| Characteristic | Total Count (n, %) | True Bleed Present<br>(TP + FN) (n, %) | Age (Mean, SD) | Mean Age When Bleed<br>Truly Present (Mean, SD) |
| --- | --- | --- | --- | --- |
| Female | 202, 50 | 31, 15.3 | 66.6, 18.3 | 70.3, 16.4 |
| Male | 202, 50 | 49, 24.3 | 60.3, 19.4 | 62.0, 15.8 |

**Table III.** Frequency of false positive outputs and assessment of cause.

| Reason | Count |
| --- | --- |
| Motion | 4 |
| Residual Contrast | 4 |
| Unidentified Reason | 4 |
| Streak | 3 |
| Dural Thickening/Calcifications (Falx/Tentorium) | 3 |
| Meningioma | 2 |
| Basal Ganglia Calcification and Streak | 1 |
| Chronic Post-Surgical Dural Thickening (Craniotomy) | 1 |
| Hemoconcentrated Blood the Dural Venous Sinuses | 1 |
| Parenchymal Calcifications | 1 |
| Volume Averaging or Hemoconcentration | 1 |

**Table IV.**
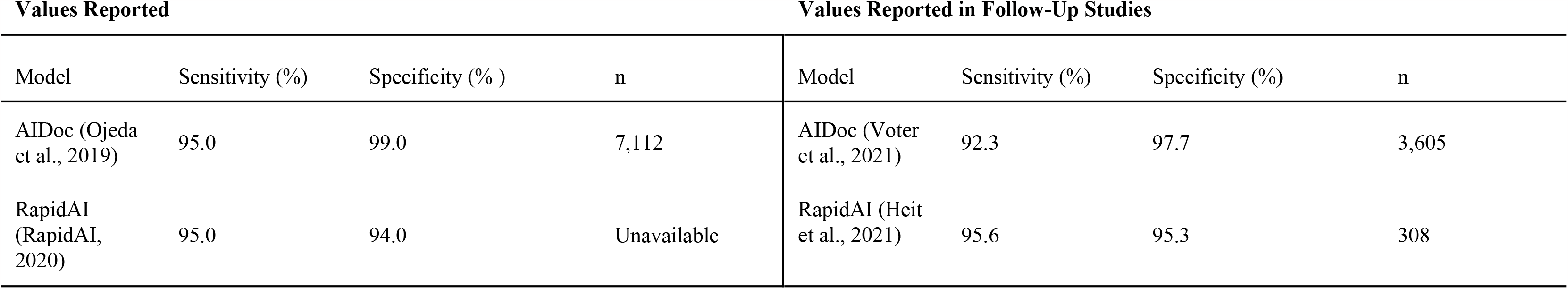
Sensitivity and specificity for two commercially available AI products for the detection of ICH and sensitivity and specificity found in follow-up studies.

Model success was evaluated by sex. The percentages of FP, FN, TP, and TN outputs for females were: 5.9%, 7.9%, 7.4%, and 78.7% (Figure I). This resulted in the following sensitivity, specificity, accuracy, PPV, and NPV: 48.4%, 95% CI [30.8%-66.0%], 93.0% [89.2%-96.8%], 86.1% [81.4%-90.9%], 55.6% [36.8%-74.3%], and 90.9% [86.6%-95.1%] (Table I).

For males, the percentages of FP, FN, TP, and TN outputs were 5.9%, 3.5%, 20.8%, and 69.8% respectively (Figure I). This resulted in the following sensitivity, specificity, accuracy, PPV, and NPV: 85.7%, 95% CI [75.9%-95.5%], 92.2% [87.9%-96.4%], 90.6% [86.6%-94.6%], 77.8% [66.7%-88.9%], and 95.3% [91.9%-98.7%] (Table I).

A chi-square test of independence was performed to assess whether correct (FP or TP) or incorrect (FN or TN) software output is dependent on sex. It was determined that model correctness is not dependent on sex: *X*^*2*^ (1 degree of freedom, *N* = 404) = 1.95, *p* = .162 (*p* > 0.05).

## Discussion

In this study of 406 cases, the diagnostic performance of Rapid ICH was retrospectively assessed. Rapid ICH exhibited a lower sensitivity and specificity at our site than advertised, although its performance was only significantly different by the measurement of sensitivity. Sensitivity is a measure of how adept a test is at identifying true cases of a condition and decreases with increasing counts of FN outputs, or, in our case, missed bleeds. Thus, institution specific analysis of Rapid’s behavior is crucial so that the software’s limitations are understood, and the model is utilized appropriately by physicians.

We identified the common etiologies associated incorrect model outputs. The model produced more FP outputs than FN outputs. The most common FP outputs produced by the software resulted from motion, residual contrast, or could not be associated with a feature. FP results are particularly significant because they add to the burden on hospitals and physicians by unnecessarily flagging cases for review. This undermines AI’s primary selling point of increased efficiency. It is pertinent to identify the features that trigger FP results, as these represent areas for improvement in the model in selecting for ICH.

It is of the utmost importance that FN outputs are identified and classified so that the algorithms for automatic ICH detection can be improved and confidently integrated into hospital workflow. We found that most FN outputs occurred on scans with SDH and IPH at our site. There is increasing concern about growing radiologist workload and how fatigue may impact error rates (Bruls & Kwee, 2020). AI models have the potential to address this concern, and given their automatic nature, even compensate for human error. But they can only do so if the FN frequency is lower than that of the overburdened radiologist. Otherwise, this software could contribute to missed bleeds and catastrophic patient outcomes when incorporated into hospital workflow.

The error rate of radiologists is incredibly low (Strub et al., 2007) and previous assessment of other ICH detection software (AIDoc) has shown that residents, even when under time pressure, outperform the algorithm (Kau et al., 2022). We found that scans presenting SDH resulted in the highest frequency of FN outputs (Figure II). Interestingly, in their assessment of misidentified ICH by residents, Strub and colleagues found that SDH was the primary bleed type incorrectly assessed by their doctors (39% of cases) (Strub et al., 2007). This points to both areas for improvement in the algorithm and avenues for further research: RapidAI could potentially compensate for the human error in SDH diagnosis specifically if the model is fine-tuned to identify SDH. As RapidAI continues to update their software, further studies comparing doctor to AI performance are warranted.

We add to the growing body of studies that assess the reported performance of AI in identifying ICH on new datasets, thus exploring the generalizability of these models. As these models become increasingly prevalent in the clinical workspace it is important to understand how well they function so that they can be appropriately utilized. It is also the responsibility of doctors and hospitals to identify shortcomings in the software so that areas of improvement can be addressed by developers. This will drive necessary change so that AI achieves its potential of improving patient outcomes while simultaneously reducing physician burden. Table IV shows the sensitivity and specificity of two commonly used commercially available AI products for the detection of ICH and the values found in subsequent studies. These statistics vary across sites. In their 2021 study, Heite and colleagues analyzed the performance of RapidAI in a study of 308 scans and found the following sensitivity, specificity, PPV, and NPV for the software: 95.6%, 95.3%, 95.6%, and 95.3%. The disparity between our findings, and other assessments of RapidAI’s performance indicate the need for further investigation of the generalizability of the model.

In another study, AIDoc was assessed and it was determined that its performance was not consistent across trials (Table IV; (Voter et al., 2021)). The performance of the model as a function of age and sex was also analyzed to determine if patient demographic factors contribute to varying model performance. In the univariable analysis but not their multivariable analysis, it was determined that model performance did in fact depend on sex. In the current study, erroneous outputs did not significantly depend on sex. This points to further investigation into the impact of patient demographic factors on the model’s behavior.

This study must be considered within the context of its limitations. A larger sample size would bolster statistical power and representativeness of the data. The confidence intervals were wide, particularly for the overall sensitivity and PPV, female sensitivity and PPV, and male sensitivity, specificity, and PPV. Future studies with more examinations would provide a robust description of the model’s performance and allow more thorough investigation of the impact of demographic factors. Moreover, this study involved scans from only one scanner model. A more rigorous analysis would involve data from multiple scanner models to determine if the software’s varying performance depends on scanner type. Lastly, Rapid ICH has since been updated to a new version with advertised improvements in sensitivity and specificity(*Rapid ICH Receives New FDA Clearance with Highest Sensitivity and Specificity on the Market*, 2022). The present study considers the previous version of the software. The performance of the software as it is updated should be considered in future studies so that physicians can understand the capabilities and shortcomings of this tool as it evolves over time.

Despite its limitations, this study is the first to quantify the performance of Rapid ICH at our site, adding to the growing body of evidence supporting independent validation of machine learning products for medical imaging at all sites in which they are utilized.

## Conclusion

In conclusion, the sensitivity Rapid ICH is not in concordance with the values published by the FDA when the software was used to identify ICH at this regional medical center. Additionally, the performance of RapidAI does not significantly vary with sex. These results underscore the essentiality in per-hospital evaluation of AI software for automatic ICH detection prior to implementation into the hospital workflow. Currently, Rapid ICH is most appropriately utilized in conjunction with a doctor.

## Data Availability

All data produced in the present study are available upon reasonable request to the authors

## Acknowledgements

The authors would like to acknowledge Vision Radiology for providing access to the case studies used for this paper.

## Declarations

### Ethics approval

Pearl IRB, an independent institutional review board, gave ethical approval for this work. All procedures were performed in accordance with the ethical standards of Pearl IRB and with the 1964 Helsinki declaration and its later amendments or comparable ethical stands.

### Informed consent

Formal informed consent was not required given the retrospective nature of the study and approval by Pearl IRB.

## References

Bruls, R., & Kwee, R. (2020). Workload for radiologists during on-call hours: dramatic increase in the past 15 years. Insights into imaging, 11(1), 1–7.

Chen, M. M., Golding, L. P., & Nicola, G. N. (2021). Who will pay for AI? Radiology: Artificial Intelligence, 3(3).

Elliott, J., & Smith, M. (2010). The acute management of intracerebral hemorrhage: a clinical review. Anesthesia & Analgesia, 110(5), 1419–1427.

Harris, S. (2019). AI in medical imaging to top $2 billion by 2023. In.

Kau, T., Ziurlys, M., Taschwer, M., Kloss-Brandstätter, A., Grabner, G., & Deutschmann, H. (2022). FDA-approved deep learning software application versus radiologists with different levels of expertise: detection of intracranial hemorrhage in a retrospective single-center study. Neuroradiology, 64(5), 981–990.

Ojeda, P., Zawaideh, M., Mossa-Basha, M., & Haynor, D. (2019). The utility of deep learning: evaluation of a convolutional neural network for detection of intracranial bleeds on non-contrast head computed tomography studies. Medical Imaging 2019: Image Processing,

Rao, B., Zohrabian, V., Cedeno, P., Saha, A., Pahade, J., & Davis, M. A. (2021). Utility of artificial intelligence tool as a prospective radiology peer reviewer—detection of unreported intracranial hemorrhage. Academic radiology, 28(1), 85–93.

Rapid ICH Receives New FDA Clearance with Highest Sensitivity and Specificity on the Market. (2022). RapidAI.

RapidAI. (2020). Rapid ICH Suspected Intracranial Hemorrhage Identification: A Technical Overview of Rapid ICH and the Role of Machine Learning. In. RapidAI: iSchemaView.

RapidAI. (2023). Rapid ICH and Rapid Hyperdensity. https://www.rapidai.com/rapid-ich-hyperdensity

Reeves, K. (2022). Times are tight: staff shortages prompt new strategies. Applied Radiology, 51(4), 27–30.

Ruutiainen, A. T., Durand, D. J., Scanlon, M. H., & Itri, J. N. (2013). Increased error rates in preliminary reports issued by radiology residents working more than 10 consecutive hours overnight. Academic radiology, 20(3), 305–311.

Strub, W. M., Leach, J. L., Tomsick, T., & Vagal, A. (2007). Overnight preliminary head CT interpretations provided by residents: locations of misidentified intracranial hemorrhage. AJNR Am J Neuroradiol, 28(9), 1679–1682. 10.3174/ajnr.A0653

Tariq, A., Purkayastha, S., Padmanaban, G. P., Krupinski, E., Trivedi, H., Banerjee, I., & Gichoya, J. W. (2020). Current clinical applications of artificial intelligence in radiology and their best supporting evidence. Journal of the American College of Radiology, 17(11), 1371–1381.

Voter, A. F., Meram, E., Garrett, J. W., & John-Paul, J. Y. (2021). Diagnostic accuracy and failure mode analysis of a deep learning algorithm for the detection of intracranial hemorrhage. Journal of the American College of Radiology, 18(8), 1143–1152.

